# Predictors and consequences of HIV status disclosure to adolescents living with HIV in Eastern Cape, South Africa

**DOI:** 10.1101/2021.11.19.21266573

**Authors:** Olanrewaju Edun, Yulia Shenderovich, Siyanai Zhou, Elona Toska, Lucy Okell, Jeffrey W Eaton, Lucie Cluver

## Abstract

**Introduction:** The World Health Organization recommends full disclosure of HIV-positive status to adolescents who acquired HIV perinatally (APHIV) by age 12. However, even among adolescents (aged 10-19) already on antiretroviral therapy (ART), disclosure rates are low. Caregivers often report the child being too young and fear of disclosure worsening adolescents’ mental health as reasons for non-disclosure. Evidence is limited about predictors of disclosure and its association with adherence, viral suppression, and mental health outcomes among adolescents in sub-Saharan Africa.

**Methods:** Analyses included three rounds (2014-2018) of data collected among a closed cohort of adolescents living with HIV in Eastern Cape, South Africa. We used logistic regression with respondent random-effects to identify factors associated with disclosure, and assess differences in ART adherence, viral suppression, and mental health symptoms between adolescents by disclosure status. We also explored differences in the change in mental health symptoms and ART adherence between study rounds and disclosure groups with logistic regression.

**Results:** 813 APHIV were interviewed at baseline, of whom 769 (94.6%) and 729 (89.7%) were interviewed at the second and third rounds, respectively. The proportion aware of their HIV-positive status increased from 63.1% at the first round to 85.5% by the third round. Older age (adjusted odds ratio (aOR): 1.24; 1.07 – 1.43) and living in an urban location (aOR: 2.76; 1.67 – 4.45) were associated with disclosure between interviews. There was no association between awareness of HIV-positive status and ART adherence, viral suppression, or mental health symptoms among all APHIV interviewed. However, among APHIV not aware of their status at baseline, adherence decreased at the second round among those who were disclosed to (N=131) and increased among those not disclosed to (N=151) (interaction aOR: 0.39; 0.19 – 0.80). There was no significant difference in the change in mental health symptoms between study rounds and disclosure groups.

**Conclusions:** Awareness of HIV-positive status was not associated with higher rates of mental health symptoms, or lower rates of viral suppression among adolescents. Disclosure was not associated with worse mental health. These findings support the recommendation for timely disclosure to APHIV, however, adherence support post-disclosure is important.

## Introduction

In 2020, there were an estimated 1.75 million adolescents (aged 10-19 years) living with HIV (ALHIV) globally, of whom nearly 90% lived in sub-Saharan Africa (SSA) [1]. Despite declines in the number of new HIV infections among adolescents, increased access to antiretroviral therapy (ART), resulting in improved survival of children with perinatally acquired HIV, has led to a growing population of adolescents with perinatally acquired HIV (APHIV) [2,3]. Recent estimates from surveys in Southern African suggest that as many two-thirds of ALHIV acquired HIV perinatally [2].

Levels of ART adherence, treatment retention and viral suppression are lower amongst APHIV compared to younger children and adults [4,5], and evidence for effective interventions to improve these outcomes is sparse [6,7]. This is likely due to the complex biological and psychosocial changes associated with adolescence, coupled with the challenges of living with HIV. In addition, APHIV experience distinct challenges related to their mode of acquisition of HIV, which further affect their treatment outcomes [5]. Children who acquired HIV perinatally are often not told their HIV-positive status at the time of diagnosis or even ART initiation, with disclosure only later in life often after adolescence. The World Health Organization (WHO) recommends that disclosure occur at school age (6-12 years), with information provided incrementally from younger ages and full disclosure by age 12 [8]. Adolescents who are aware of their HIV-positive status have been found to report better ART adherence [9], be more likely to be retained on treatment and more easily access social support compared to those unaware [10–12]. They have also been shown to be more likely to engage in safer sex and have lower rates of mortality [11,13].

However, a recent systematic review found that the proportion of APHIV who have their HIV diagnosis disclosed to them ranged widely from 9% to 72% [14]. Among APHIV, prevalence of disclosure increased with age [12,14,15]. Other factors associated with disclosure include female gender, imminent onset of sexual debut, awareness of caregiver’s HIV-positive status, and higher levels of caregiver education [12,16–18]. In contrast, reasons reported by caregivers for non-disclosure include the child’s young age, fear of disclosure’s negative mental health consequences, risk of stigma if parents’ status is unmasked by the child, and a lack of clear guidelines on disclosure [9,10,14,15,19,20]. Identifying the predictors of disclosure to APHIV is essential to distinguish sub-groups of APHIV who may be at increased risk of non-disclosure and require disclosure-focused interventions.

Currently, data are limited on the association between HIV-status disclosure and virologic outcomes, with most studies evaluating the relationship between disclosure and ART adherence [9,21–23]. There is also limited evidence on changes in adherence post-disclosure with most studies assessing cross-sectional relationships between disclosure and adherence. Qualitative studies have reported adolescents experiencing shock and sadness post-disclosure [12,24], however, it is unclear if this impacts ART adherence.

Studies on the effects of disclosure on mental health outcomes amongst APHIV in SSA have reported conflicting results. Menon *et al*. found that, among 127 adolescents (11-15 years) on ART in Zambia, those who had not been disclosed to had higher levels of emotional difficulties [25], whereas Vreeman *et al*. in 2014 found that among 792 children and adolescents (6-14 years) in Western Kenya, rates of depression were higher among those disclosed to [26]. More recently, an intervention to increase rates of disclosure in Kenya which included 285 adolescents (10-14 years) also found higher rates of depression in the intervention (disclosed) arm after six months of starting the disclosure intervention [27]. However, there was no difference at 12, 18 and 24 months [27]. Due to the paucity of evidence from studies conducted in SSA, current WHO disclosure guidelines were largely informed by studies from high-income countries [8]. These studies, mostly conducted in the USA found lower levels of depression [28,29], anxiety [29,30] and psychological adjustment problems [31,32] among children or adolescents aware of their HIV-positive status, compared to those unaware. Though encouraging, these findings may not be generalizable in SSA where levels of stigma may be higher and access to mental health support lower.

We analysed data from a community-traced cohort study in Eastern Cape, South Africa to 1) identify factors associated with disclosure longitudinally, 2) assess the difference in self-reported ART adherence, viral suppression, and mental health symptoms by disclosure status, and 3) explore changes between study rounds in mental health symptoms and ART adherence by disclosure status. These will improve our understanding of this important process for caregivers and healthcare providers and determine need for adherence and psychological support during and after the disclosure process.

## Methods

### Study population

The study traced all adolescents living with HIV (aged 10-19 at baseline) who had initiated ART from all 52 ART clinics in a large urban, peri-urban, and rural district in Eastern Cape, South Africa [33]. Adolescents were identified via paper and computerized records and traced to their homes. At baseline (2014-2015), 1046 ALHIV were recruited, representing 90% of the 1176 patient records identified. These ALHIV were followed-up over a 4-year period (2014-2018) for three rounds of data collection. Quantitative interviews were self-administered using standardised questionnaires on tablet devices. Available viral load data were extracted from participants’ clinical records and linked to their questionnaire data [33,34].

This analysis was restricted to adolescents who acquired HIV perinatally (APHIV) (Figure 1) determined by ART initiation age 10-years or younger and validated using other supporting evidence such as history of parental death, maternal HIV-status and self-reported sexual history [33,35].

**Figure 1.**
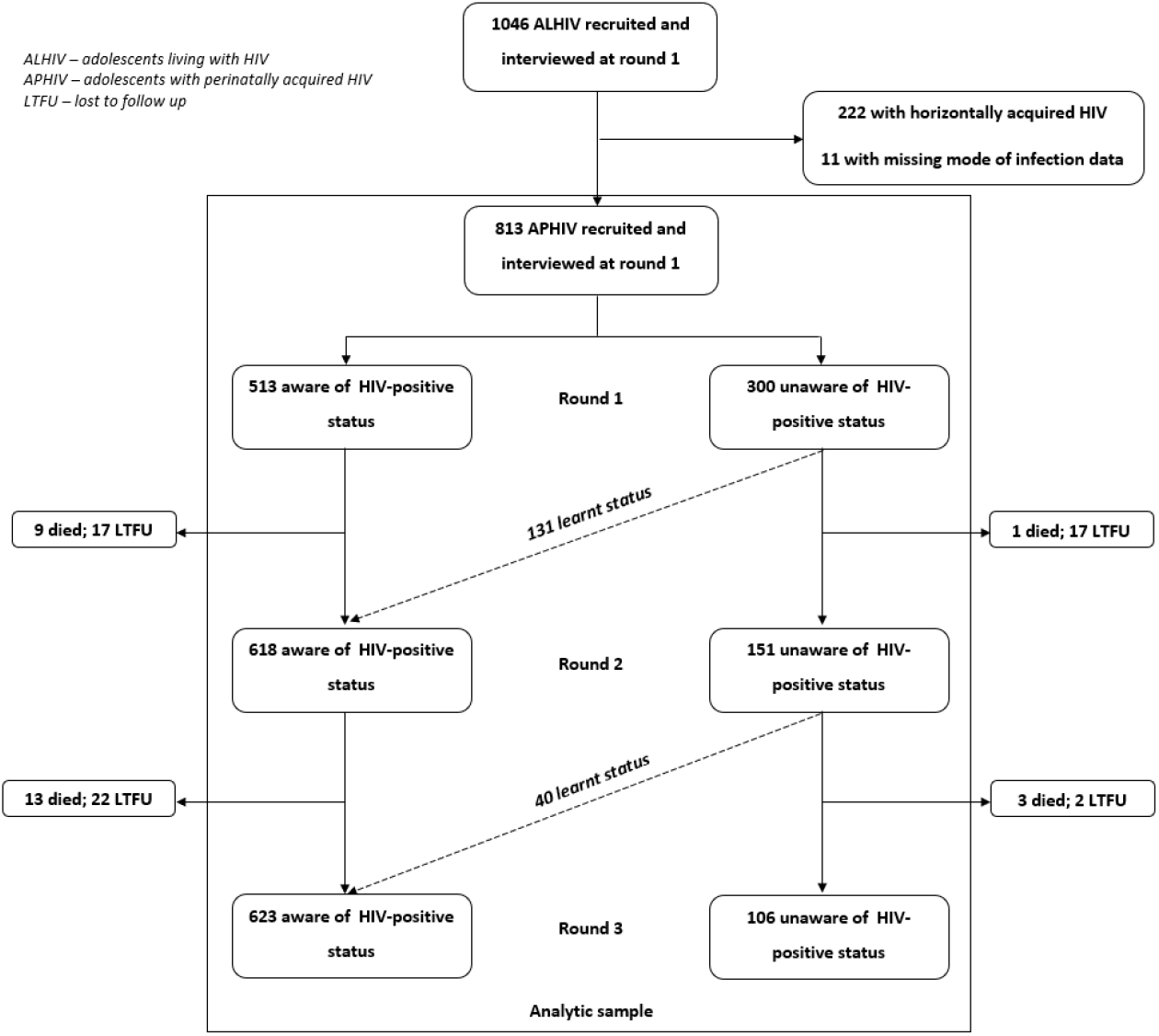
Flow diagram of study and analytic sample.

### Study measures

#### Outcome variables

The outcome for the first objective was learning one’s HIV-positive status (disclosure to the adolescent). Mental health symptomology (anxiety, depression, and suicidality), and HIV-treatment outcomes (ART adherence and viral load suppression) were the outcomes for the second and third objectives.

At baseline, awareness of HIV-status was assessed firstly through healthcare worker report, then with primary caregivers during the consent process. In cases of discrepancies, interviewers asked adolescents if they knew what their illness was, if they had ever tested for HIV and if they knew what their medication was [36]. For APHIV unaware at baseline, awareness was reassessed at subsequent study rounds from primary caregivers and adolescents during the consent process. Adolescents unaware of their HIV-positive status were asked about “illness” and “medication” as opposed to “HIV” and “ART” in study questionnaires. Disclosure was defined as being unaware of one’s HIV-positive status at baseline or the second study round and being aware at the subsequent study round.

Anxiety symptoms in the past month were assessed using a 14-item abbreviated version of the Children’s Manifest Anxiety Scale-Revised [37]. This scale, which has previously been validated in studies among AIDS-affected children [38], included “no” and “yes” responses to the experience of each symptom, coded as “0” and “1” with a total score range of 0-14. Depression symptoms in the past two weeks were assessed using the Child Depression Inventory (short form) 10-item version [39]. This scale, which has also been used and validated in other SSA populations [40–43], had a 3-point Likert-type scale ranging from 0-2 with a total score range of 0-20. Suicidality symptoms in the past months were assessed using the Mini International Psychiatric Interview for Children and Adolescents suicidality and self-harm subscale [44]. This 5-item scale which has been validated in developed world populations and adapted in sub-Saharan African settings [45–47], included “no” and “yes” responses to the experience of each symptoms, coded as “0” and “1” with a total score range of 0-5. All symptoms in these measures had equal weight. Due to the small number of participants endorsing the most severe symptoms, we created binary variables for any symptoms vs. none on each of three scales.

An adapted version of the standardised Patient Medication Adherence Questionnaire was used to assess self-reported ART adherence in the past week, alongside measures developed in Botswana [48,49]. Adherence was defined by reporting currently taking ART and not having missed any doses in the past seven days (including weekdays and weekend) [34]. We included viral load results within a one-year period before or after their questionnaire interview dates for the respective study rounds. Viral suppression was defined as viral load <1000 copies/ml.

### Explanatory and control variables

The main explanatory variable of interest for the mental health and HIV-treatment outcomes was awareness of HIV-positive status, as described above. Other control variables were sociodemographic information— age, sex, dwelling type, orphanhood status, relationship with primary caregiver and household poverty. Household poverty was assessed by measuring access to the top eight socially-perceived necessities for children as defined by the Centre for South African Social Policy [50]. Adolescents were classified as living in poverty if they reported not having access to all eight necessities. We also included measures of abuse (physical and emotional) and stigma (anticipated and secondary) as control variables. Physical and emotional abuse were measured using items from the UNICEF Measures for National-level Monitoring of Orphans and Vulnerable Children [51]. Anticipated stigma was measured using two items from the ALHIV-Stigma Scale which assessed adolescents’ views of the community’s perception towards HIV and has been used previously among ALHIV in SSA [52]. Secondary stigma due to HIV in families or households was measured using the 6-item Stigma-By-Association scale which has been validated for use in South Africa [53]. Adolescents were categorized as having experienced physical or emotional abuse and anticipated or secondary stigma if they self-reported at least one experience of these in the past year.

### Data analyses

Characteristics of study participants overall and by awareness of HIV-status in each round were summarized using means, standard deviations, median, interquartile ranges, and proportions. Differences between participant characteristics by awareness of HIV-status at all study rounds were calculated using t-tests for continuous variables and chi-square tests for categorical variables.

Second, among APHIV who were unaware of their HIV-status at rounds one and two, we used random-intercepts logistic regression to identify factors associated with disclosure between rounds. The outcome was learning one’s HIV-status at round two or three and the explanatory variables were demographic, psychological, and social factors at the survey prior to disclosure. Individual-level random intercepts were used to account for the repeated observations of the same individuals. Variables identified *a priori* to be associated with disclosure in the literature such as age, sex, dwelling location, caregiver relationship and orphanhood status [12,14,15,54], and the study round, were included in a multivariate regression model.

Third, to assess if awareness of HIV-positive status was associated with self-reported ART adherence, viral suppression and mental health problems symptoms, data for all APHIV who were interviewed at any of the three rounds were analysed. Logistic regression with individual-level random intercepts was used to estimate the odds of ART adherence, viral suppression and reporting any symptom of depression, anxiety, and suicidality. We adjusted for potential confounders identified from our conceptual framework and the study round in a multivariate random-intercepts logistic regression model. A sensitivity analyses was conducted using only those interviewed at all three study rounds.

Lastly, again among those who were unaware of their HIV-status at rounds one and two, we analysed whether there was a differential change in reporting any mental health problems symptom or ART adherence between study rounds for APHIV who were disclosed to versus those who were not. Change in viral suppression was not examined due to limited availability of viral load data. We specified the following logistic regression model to estimate if the odds of reporting any anxiety, depression or suicidality symptom, or ART adherence between study rounds (rounds one to two and two to three), was different between those who learnt their HIV-status and those who did not:

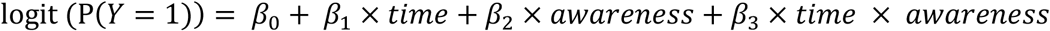

Where *Y* represents our mental health outcomes or ART adherence, *time* is a dummy variable indicating round one or two, and *awareness* is a dummy variable indicating awareness of status at round two. *β*_3_ indicates the difference between the log-odds ratio comparing round one vs. two in those who learnt their status at two and the log-odds ratio comparing round one vs. two in those who did not. We reported exponentiated *β*_3_ estimates (both crude and adjusted for factors hypothesized to be associated with mental health symptoms and ART adherence from our conceptual framework (Figure S1)). We considered p-values ≤0.05 as statistically significant and all p-values are two-sided. Analyses were conducted in R version 3.6.1 [55].

### Ethical approval

Ethical approval for the study was granted by the Institutional Review Boards at the Universities of Cape Town (CSSR 2013/4) and Oxford (SSD/CUREC2/12-21). Provincial approval was obtained from the Eastern Cape Departments of Education and Health and participating health facilities to conduct the study and access medical records. Written informed consent for the interviews and to access clinical records was obtained from participants and their primary caregivers. Ethical approval for the secondary analyses of study data was obtained from the Imperial College Research Governance and Integrity Team (20IC6451).

## Results

### Summary of participant characteristics

Of 1046 ALHIV recruited and interviewed at baseline, 813 (77.7%) acquired HIV perinatally (APHIV). Our analyses included the 813 APHIV interviewed at baseline, and 769 (94.6%) and 729 (89.7%) interviewed at the second and third rounds respectively. 729 were interviewed at all three rounds. The median age of eligible participants was 13-, 14- and 15-years at rounds one, two and three, respectively. The proportion of males and females were nearly equal at all study rounds (Table 1). The proportion of APHIV who were aware of their status increased from 63.1% at baseline to 80.4% at round two and 85.5% at round three. The proportion of APHIV reporting any symptom of depression, anxiety and suicidality reduced in successive study rounds.

**Table 1.** Summary of participant characteristics at all three study rounds.

|  | Study round |  |  |
| --- | --- | --- | --- |
|  | 1<br>(N = 813 ) | 2<br>(N = 769) | 3<br>(N = 729) |
| Age in years, median (IQR) | 13 (11 – 14) | 14 (12 – 16) | 15 (13 – 17) |
| Female, n (%) | 410 (50.4) | 390 (50.7) | 369 (50.6) |
| Aware of HIV status, n (%) | 513 (63.1) | 618 (80.4) | 623 (85.5) |
| <sup>†</sup> Urban dwelling, n (%) | 611 (75.2) | 581 (75.6) | 556 (76.3) |
| Caregiver biological parent, n (%) | 348 (42.8) | 296 (38.5) | 277 (38.0) |
| Household poverty, n (%) | 531 (65.3) | 589 (76.6) | 484 (66.4) |
| Any parental loss, n (%) | 502 (61.7) | 485 (63.1) | 517 (70.9) |
| Any emotional abuse in last year, n (%) | 202 (24.8) | 201 (26.1) | 138 (18.9) |
| Any physical abuse in last year, n (%) | 261 (32.1) | 207 (26.9) | 110 (15.1) |
| Any anticipated stigma reported, n (%) | 198 (24.4) | 155 (20.2) | 115 (15.8) |
| Any secondary stigma in last year, n (%) | 150 (18.5) | 63 (8.2) | 27 (3.7) |
| Self-reported past week ART adherence, n (%) | 559 (68.8) | 517 (67.2) | 568 (77.9) |
| <sup>‡</sup> Viral suppression (using results one year before, or after interview), n (%) | 400 (78.7) | 360 (77.3) | 190 (66.2) |
| Any depression symptom in past two weeks, n (%) | 352 (43.3) | 260 (33.8) | 205 (28.1) |
| Any anxiety symptom in past month, n (%) | 505 (62.1) | 218 (28.3) | 183 (25.1) |
| Any suicidality risk in past month, n (%) | 46 (5.7) | 24 (3.1) | 24 (3.3) |

<sup>†</sup> Type of dwelling data missing for 1 (0.1%) adolescent at round 2.
<sup>‡</sup> Only 508 (62.5%), 466 (60.6%) and 287 (39.4%) adolescents had viral load results one year before or after their round one, two and three interviews respectively.

At baseline, APHIV who lived in urban locations and had experienced the loss of a parent at were more likely to be aware of their HIV-positive status (Table 2). The mean age amongst those aware (13.6 years) was older compared to those unaware of their status (11.5 years). Other participant characteristics by awareness of HIV-positive status and study round can be found in Table 2 and Table S1.

**Table 2.**
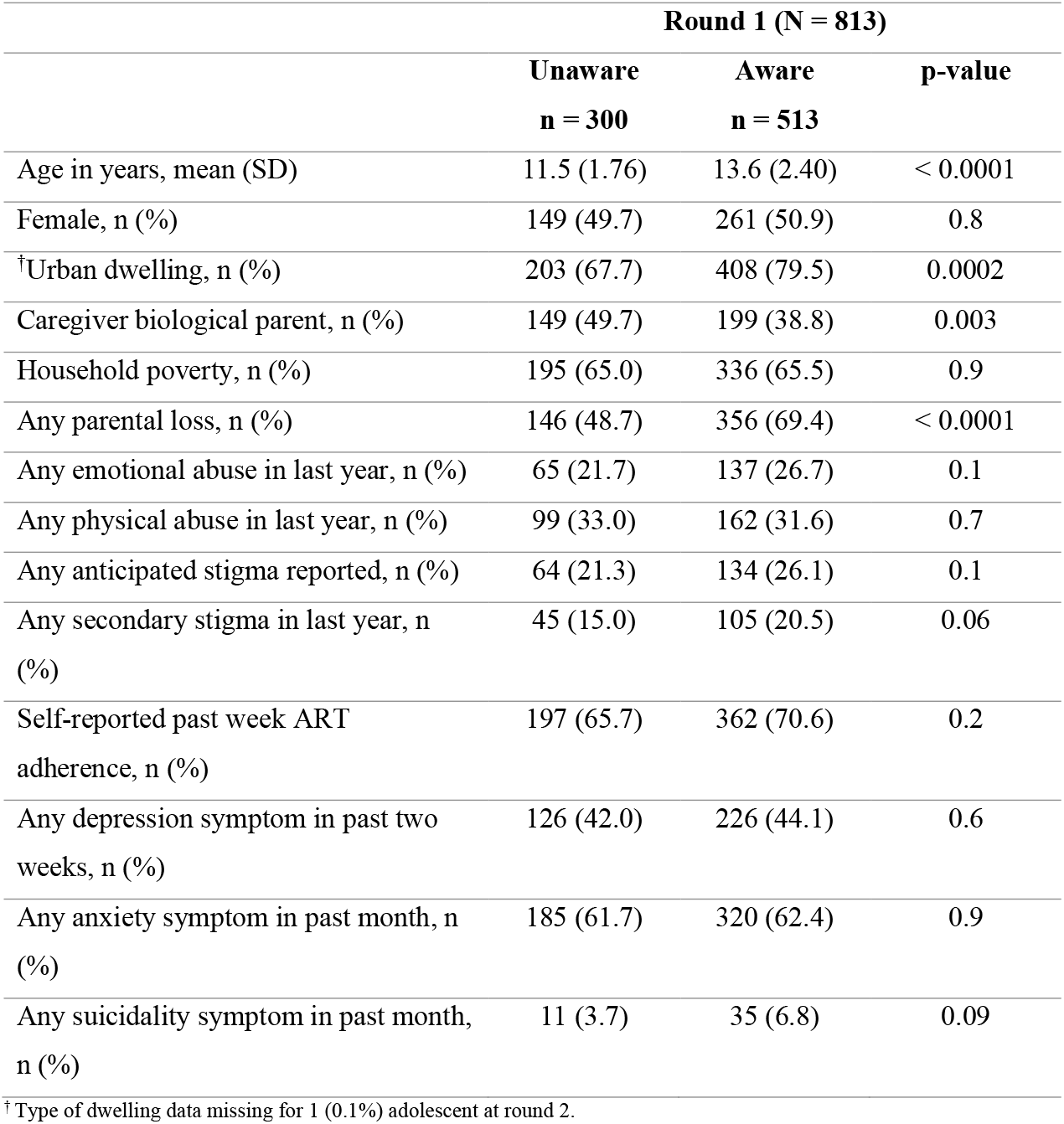
Baseline characteristics of study participants stratified by awareness of their HIV-positive status. See supplementary material Table S1 for full table including characteristics at rounds two and three.

|  | Round 1 (N = 813) |  |  |
| --- | --- | --- | --- |
|  | Unaware<br>n = 300 | Aware<br>n = 513 | p-value |
| Age in years, mean (SD) | 11.5 (1.76) | 13.6 (2.40) | < 0.0001 |
| Female, n (%) | 149 (49.7) | 261 (50.9) | 0.8 |
| <sup>†</sup>Urban dwelling, n (%) | 203 (67.7) | 408 (79.5) | 0.0002 |
| Caregiver biological parent, n (%) | 149 (49.7) | 199 (38.8) | 0.003 |
| Household poverty, n (%) | 195 (65.0) | 336 (65.5) | 0.9 |
| Any parental loss, n (%) | 146 (48.7) | 356 (69.4) | < 0.0001 |
| Any emotional abuse in last year, n (%) | 65 (21.7) | 137 (26.7) | 0.1 |
| Any physical abuse in last year, n (%) | 99 (33.0) | 162 (31.6) | 0.7 |
| Any anticipated stigma reported, n (%) | 64 (21.3) | 134 (26.1) | 0.1 |
| Any secondary stigma in last year, n (%) | 45 (15.0) | 105 (20.5) | 0.06 |
| Self-reported past week ART adherence, n (%) | 197 (65.7) | 362 (70.6) | 0.2 |
| Any depression symptom in past two weeks, n (%) | 126 (42.0) | 226 (44.1) | 0.6 |
| Any anxiety symptom in past month, n (%) | 185 (61.7) | 320 (62.4) | 0.9 |
| Any suicidality symptom in past month, n (%) | 11 (3.7) | 35 (6.8) | 0.09 |

<sup>†</sup> Type of dwelling data missing for 1 (0.1%) adolescent at round 2.

### Predictors of disclosure

Among the 300 APHIV unaware of their status at baseline, 282 participated in round two. Among these 282 adolescents, 131 (46.5%) became aware of their status by round two. Among the 151 adolescents who remained unaware of their status by round two, 146 participated in round three, with 40 (27.4%) learning their status. In total, 171 APHIV learnt their status between study rounds. Factors associated with learning one’s HIV-positive status were older age (adjusted odds ratio [aOR]: 1.24, 95% confidence interval [95%CI]: 1.07 – 1.44) and residing in an urban area (aOR: 2.83, 95%CI: 1.72 – 4.67) at the previous time point (Table 3).

**Table 3.** Random-intercepts logistic regression results showing odds of learning HIV-positive status (disclosure) between study rounds 1-2 and 2-3 by demographic and psychosocial factors at the survey prior to disclosure. See Table S2 for characteristics stratified by rounds.

| Baseline characteristics | Disclosure status |  | Odds ratio<br>(95% CI) | Adjusted odds ratio<br>(95% CI) |
| --- | --- | --- | --- | --- |
|  | mean (SD) or n (%) |  |  |  |
|  | No disclosure<br>n = 111 | Disclosure<br>n = 171 |  |  |
| N = 282 |  |  |  |  |
| Age | 11.2 (1.45) | 11.7 (1.84) | 1.19 (1.00 – 1.42)* | 1.24 (1.07 – 1.44)** |
| Female | 54 (48.6) | 88 (51.5) | 1.23 (0.76 – 2.01) | 1.19 (0.78 – 1.82) |
| Urban dwelling | 57 (51.4) | 130 (76.0) | 3.04 (1.72 – 5.36)*** | 2.83 (1.71– 4.67)*** |
| Caregiver is biological parent | 58 (52.3) | 80 (46.8) | 0.96 (0.60 – 1.54) | 0.85 (0.54 – 1.37) |
| Household poverty | 77 (69.4) | 110 (64.3) | 0.72 (0.43 – 1.18) | 0.81 (0.51 – 1.30) |
| Any parental loss | 53 (47.7) | 85 (49.7) | 1.50 (0.91 – 2.47) | 1.11 (0.69 – 1.81) |
| Any emotional abuse in last year | 15 (13.5) | 44 (25.7) | 1.66 (0.94 – 2.92) | 1.21 (0.71 – 2.06) |
| Any physical abuse in last year | 35 (31.5) | 58 (33.9) | 1.10 (0.66 – 1.81) | 0.89 (0.56 – 1.42) |
| Any anticipated stigma reported | 22 (19.8) | 36 (21.1) | 1.38 (0.77 – 2.48) | 1.15 (0.68 – 1.94) |
| Any secondary stigma in last year | 13 (11.7) | 29 (17.0) | 1.20 (0.60 – 2.43) | 1.00 (0.53 – 1.89) |
| Any depression symptom | 40 (36.0) | 76 (44.4) | 1.11 (0.69 – 1.79) | 0.93 (0.59 – 1.45) |
| Any anxiety symptom | 66 (59.5) | 109 (63.7) | 1.15 (0.72 – 1.82) | 0.80 (0.50 – 1.30) |
| Any suicidality risk | 1 (0.9) | 10 (5.8) | 2.60 (0.62 – 10.95) | 1.80 (0.46 – 7.10) |

\*\*\*p ≤ 0.001, \*\*p ≤ 0.01, \*p ≤ 0.05; All models were adjusted for age, sex, dwelling type, caregiver relationship, orphanhood status, anticipated and secondary stigma, and study round.

### Association between awareness of HIV-positive status and ART adherence, viral suppression, and mental health symptoms

Among APHIV interviewed at any of the three study rounds, including those who were already aware of their HIV-status before the start of the study, there was no association between awareness of HIV-positive status and ART adherence, regardless of the time of disclosure (Table 4).

**Table 4.** Mixed effects logistic regression results showing crude and adjusted odds ratio and 95% confidence intervals (CI) for factors associated with self-reporting past week ART adherence and having a suppressed viral load within a year before or after interview.

|  | Self-reported past week ART adherence<br>(N = 813, 769 and 729 at rounds 1, 2 and 3) |  | Viral suppression<br>(N = 508, 466 and 287 at rounds 1, 2 and 3) |  |
| --- | --- | --- | --- | --- |
|  | Odds ratio<br>(95% CI) | Adjusted odds ratio<br>(95% CI) | Odds ratio<br>(95% CI) | Adjusted odds ratio<br>(95% CI) |
| **Explanatory variable** |  |  |  |  |
| Aware of HIV status | 1.05 (0.83 – 1.32) | 1.06 (0.81 – 1.38) | 0.27 (0.09 – 0.80)* | 0.69 (0.20 – 2.44) |
| **Control variables** |  |  |  |  |
| Age | 1.00 (0.97 – 1.04) | 0.96 (0.92 – 1.01) | 0.66 (0.53 – 0.81)*** | 0.89 (0.68 – 1.18) |
| Female | 1.05 (0.86 – 1.29) | 1.06 (0.86 – 1.30) | 0.98 (0.33 – 2.93) | 0.90 (0.25 – 3.23) |
| Urban dwelling | 1.00 (0.79 – 1.27) | 1.09 (0.86 – 1.40) | 0.58 (0.17 – 2.01) | 0.57 (0.13 – 2.44) |
| Caregiver is biological parent | 0.82 (0.67 – 1.00) | 0.81 (0.64 – 1.03) | 1.21 (0.53 – 2.76) | 1.24 (0.44 – 3.52) |
| Household poverty | 0.96 (0.78 – 1.18) | 0.93 (0.75 – 1.16) | 0.43 (0.20 – 0.95) | 0.35 (0.14 – 0.89)* |
| Any parental loss | 1.14 (0.92 – 1.40) | 1.05 (0.82 – 1.35) | 0.74 (0.29 – 1.91) | 1.66 (0.49 – 5.64) |
| Any emotional abuse in last year | 0.51 (0.41 – 0.63)*** | 0.63 (0.49 – 0.81)*** | 2.23 (0.89 – 5.58) | 2.52 (0.80 – 7.88) |
| Any physical abuse in last year | 0.55 (0.44 – 0.68)*** | 0.69 (0.55 – 0.88)** | 2.18 (0.93 – 5.07) | 1.08 (0.39 – 3.04) |
| Any anticipated stigma reported | 0.71 (0.56 – 0.89)** | 0.84 (0.66 – 1.07) | 2.69 (1.02 – 7.17)* | 1.86 (0.63 – 5.51) |
| Any secondary stigma in last year | 0.58 (0.43 – 0.78)*** | 0.77 (0.56 – 1.07) | 0.97 (0.33 – 2.79) | 0.38 (0.11 – 1.36) |
| Round 2 | 0.93 (0.74 – 1.15) | 0.92 (0.73 – 1.17) | 0.60 (0.27 – 1.32) | 0.83 (0.33 – 2.10) |
| Round 3 | 1.65 (1.30 – 2.10)*** | 1.54 (1.17 – 2.03)** | 0.09 (0.04 – 0.21)*** | 0.11 (0.03 – 0.36)*** |

\*\*\*p ≤ 0.001, \*\*p ≤ 0.01, \*p ≤ 0.05

Among all APHIV interviewed, 62.5% (508 of 813), 60.1% (468 of 779) and 39.0% (292 of 749) had a viral load result within a one-year period before or after their first, second and third round interview dates, respectively. Adolescents without viral load results were older, however, there was no difference in the proportion with viral load results by awareness of status or sex (Table S3). Among those with viral load results, there was no association between awareness of HIV-positive status and viral suppression, regardless of the time of disclosure (Table 4).

There was also no significant association between awareness of HIV-positive status and the odds of reporting a symptom of anxiety, depression, or suicidality among APHIV interviewed at any of the three rounds (Table 5). Conclusions were unchanged in the sensitivity analysis restricted to respondents who participated in all three study rounds (Tables S4 and S5).

**Table 5.** Random-intercepts logistic regression results showing crude and adjusted odds ratios (OR) and 95% confidence intervals for factors associated with self-reporting a symptom of anxiety within the past month, depression within the past two weeks and suicidal ideation within the past month of study interview.

|  | Anxiety |  | Depression |  | Suicidality |  |
| --- | --- | --- | --- | --- | --- | --- |
| N = 813, 769 and 729<br>at rounds 1, 2 and 3 | Odds ratio<br>(95% CI) | Adjusted odds<br>ratio<br>(95% CI) | Odds ratio<br>(95% CI) | Adjusted odds<br>ratio<br>(95% CI) | Odds ratio<br>(95% CI) | Adjusted odds<br>ratio<br>(95% CI) |
| **Explanatory variable** |  |  |  |  |  |  |
| Aware of HIV status | 0.81 (0.66 – 0.98)* | 1.05 (0.81 – 1.36) | 0.93 (0.76 – 1.15) | 0.88 (0.69 – 1.11) | 1.16 (0.44 – 3.06) | 1.62 (0.49 – 5.37) |
| **Control variables** |  |  |  |  |  |  |
| Age | 0.93 (0.89 – 0.96)*** | 1.02 (0.97 – 1.06) | 1.02 (0.98 – 1.05) | 1.07 (1.02 – 1.11)** | 0.98 (0.82 – 1.17) | 1.25 (1.00 – 1.56)* |
| Female | 1.14 (0.96 – 1.34) | 1.19 (0.98 – 1.46) | 1.00 (0.83 – 1.19) | 0.99 (0.83 – 1.19) | 1.62 (0.58 – 4.54) | 1.63 (0.56 – 4.74) |
| Urban dwelling | 1.04 (0.86 – 1.27) | 0.94 (0.74 – 1.19) | 1.05 (0.85 – 1.30) | 1.00 (0.81 – 1.24) | 1.34 (0.43 – 4.15) | 0.82 (0.26 – 2.57) |
| Caregiver is biological<br>parent | 1.04 (0.88 – 1.23) | 0.95 (0.76 – 1.19) | 0.93 (0.78 – 1.12) | 0.93 (0.75 – 1.14) | 0.75 (0.31 – 1.84) | 0.68 (0.24 – 1.93) |
| Household poverty | 0.83 (0.69 – 0.99)* | 0.94 (0.76 – 1.16) | 1.00 (0.83 – 1.21) | 1.03 (0.85 – 1.25) | 1.47 (0.69 – 3.16) | 1.39 (0.62 – 3.12) |
| Any parental loss | 0.94 (0.79 – 1.12) | 0.95 (0.75 – 1.21) | 1.03 (0.85 – 1.24) | 0.95 (0.76 – 1.18) | 0.88 (0.34 – 2.26) | 0.75 (0.24 – 2.32) |
| Any emotional abuse in last year | 2.90 (2.38 – 3.53)*** | 2.29 (1.79 – 2.92)*** | 2.29 (1.87 – 2.82)*** | 1.83 (1.46 – 2.30)*** | 5.54 (2.74 – 11.22)*** | 2.54 (1.10 – 5.89)* |
| Any physical abuse in last year | 2.65 (2.18 – 3.24)*** | 1.80 (1.42 – 2.29)*** | 1.61 (1.32 – 1.97)*** | 1.13 (0.90 – 1.41) | 4.66 (2.27 – 9.57)*** | 2.49 (1.06 – 5.85)* |
| Any anticipated stigma reported | 1.92 (1.57 – 2.36)*** | 1.43 (1.12 – 1.82)** | 1.79 (1.44 – 2.21)*** | 1.37 (1.10 – 1.71)** | 5.05 (2.39 – 10.70)*** | 2.73 (1.24 – 6.04)* |
| Any secondary stigma in last year | 5.56 (4.09 – 7.55)*** | 3.22 (2.29 – 4.55)*** | 3.53 (2.66 – 4.67)*** | 2.46 (1.82 – 3.31)*** | 12.73 (5.56 – 29.92)*** | 6.58 (2.74 – 15.84)*** |
| Round 2 | 0.22 (0.18 – 0.28)*** | 0.22 (0.17 – 0.28)*** | 0.66 (0.54 – 0.81)*** | 0.67 (0.53 – 0.84)*** | 0.30 (0.14 – 0.63)*** | 0.28 (0.12 – 0.65)** |
| Round 3 | 0.19 (0.15 – 0.24)*** | 0.22 (0.17 – 0.29)*** | 0.50 (0.40 – 0.62)*** | 0.54 (0.42 – 0.69)*** | 0.35 (0.17 – 0.73)** | 0.39 (0.15 – 1.06) |

\*\*\*p ≤ 0.001, \*\*p ≤ 0.01, \*p ≤ 0.05

### Differential change in reporting any mental health symptom or ART adherence between study rounds for participants who were disclosed to versus those who were not

Among adolescents not aware of their status at baseline, adherence decreased at the second round among those who were disclosed to (N =131) and increased among those not disclosed to (N =151) (interaction aOR: 0.39, 95%CI: 0.19 – 0.80). However, there was no significant difference in the odds of reporting any symptom of anxiety, depression, and suicidality between rounds one and two by disclosure groups (Table 6). Among the adolescents (N =40) who learnt their status between rounds two and three, there was no significant difference in the odds of reporting any mental health symptoms or ART adherence between rounds two and three by disclosure groups (Table 7).

**Table 6.** Differential change in the odds of reporting any symptom of anxiety, depression and suicidality or ART adherence between rounds 1 (R1) and 2 (R2), between those who became aware of their HIV status between surveys, versus those remaining unaware of their status.

|  | n (%) |  |  |  | †Differential change estimate (95%CI) |  |
| --- | --- | --- | --- | --- | --- | --- |
|  | No disclosure R1-R2<br>(n = 151) |  | Disclosure R1-R2<br>(n = 131) |  | Crude | ‡Adjusted |
|  | Round 1 | Round 2 | Round 1 | Round 2 |  |  |
| **Anxiety** | 95<br>(62.9) | 30<br>(19.9) | 80<br>(61.1) | 41<br>(31.3) | 1.99 (0.96 – 4.13) | 2.10 (0.99 – 4.48) |
| **Depression** | 58<br>(38.4) | 43<br>(28.5) | 58<br>(44.3) | 44<br>(33.6) | 1.00 (0.50 – 2.00) | 0.99 (0.48 – 2.02) |
| **§Suicidality** | 4<br>(2.6) | 0<br>(0.0) | 7<br>(5.3) | 5<br>(3.8) | - | - |
| **ART adherence** | 94<br>(62.2) | 107<br>(70.9) | 92<br>(70.2) | 77<br>(58.8) | 0.41 (0.20 – 0.83)* | 0.39 (0.19 – 0.80)* |

\*\*\*p ≤ 0.001, \*\*p ≤ 0.01, \*p ≤ 0.05. †The differential change estimate represents the ratio of the odds ratios of reporting any symptom of anxiety, depression and suicidality or ART adherence between rounds 1 and 2 between those who became aware vs. those remaining unaware.
‡Adjusted for age, sex, dwelling location, household poverty, caregiver relationship, orphanhood status, physical and emotional abuse, and anticipated and secondary stigma.
§Model for suicidality failed to converge due to zero value in cell.

**Table 7.**
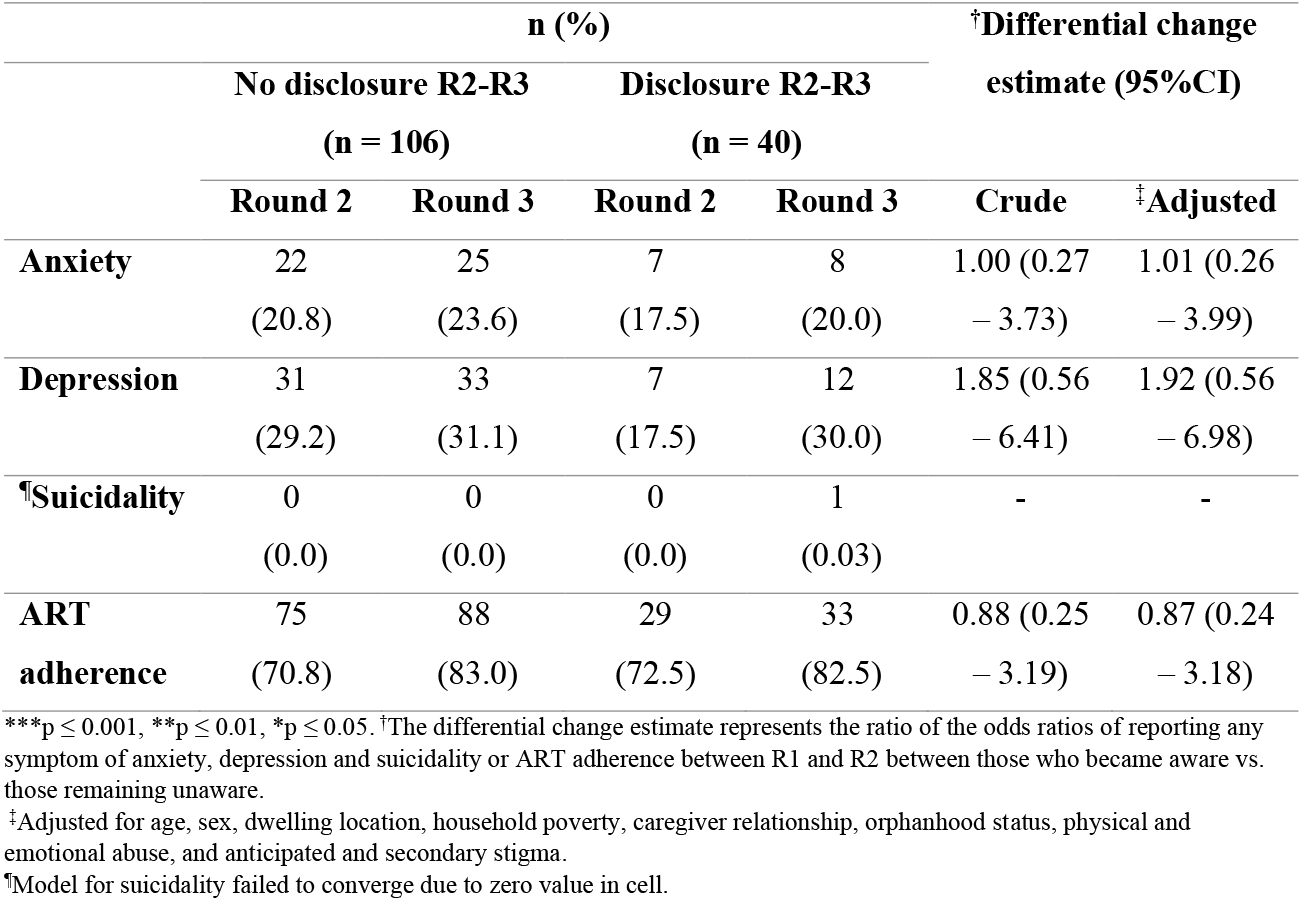
Differential change in the odds of reporting any symptom of anxiety, depression and suicidality or ART adherence between rounds 2 (R2) and 3 (R3), between those who became aware of their HIV status between surveys, versus those remaining unaware of their status.

## Discussion

Despite WHO recommendations of full disclosure by 12-years of age [8], over one-third (37%) of adolescents with perinatally-acquired HIV in this study were unaware of their HIV-positive status at baseline (22% of those ≥12-years). But 61% of those unaware learnt their status during the 4-year follow-up period. Older adolescents and those residing in an urban location were more likely to learn their HIV-positive status. Reporting of any poor mental health symptom declined over the cohort rounds and there was no significant difference in the change in mental health symptoms between adolescents who became aware of their status and those who remained unaware. Adolescents who learnt their status between baseline and the second round were less likely to report adherence post-disclosure compared to those who remained unaware. However, there was no association between awareness of HIV-positive status and ART adherence, viral suppression or symptoms of anxiety, depression, or suicidality among all APHIV during the study.

The association between age and disclosure in this study is consistent with results from other studies [12,14,15]. Also, our finding of higher odds of disclosure among urban dwelling adolescents is similar to results from a recent study among ALHIV in Tanzania [54]. This may be due greater awareness of HIV in urban communities and better access to disclosure information in urban health facilities which may facilitate disclosure.

The lack of evidence of a difference in mental health symptoms between disclosure groups is consistent with results from Vreeman *et al*., who found no difference in rates of depression at 12, 18 and 24 months between disclosure groups in a disclosure intervention in Kenya [27]. These findings may suggest that the emotional and psychological challenges experienced post-disclosure, often a concern to caregivers, may be short-lived [12,28]. However, disclosure between surveys in this study was associated with non-adherence in the subsequent survey, but only between rounds one and two. This is similar to results from a recent study among APHIV in Botswana, which found significant declines in ART adherence post-disclosure compared to pre-disclosure, although, the study lacked a non-disclosure comparison group [21]. Non-adherence post-disclosure may be due to the experience of fear, shock, and withdrawal post-disclosure [12,24] or disclosure may have been precipitated by developing adherence concerns [20]. These findings contrast with positive associations between disclosure and ART adherence previously reported by cross-sectional studies [9,56] and highlights the potential need for adherence support post-disclosure.

However, it is unclear if non-adherence post-disclosure persists. We found no association between awareness of HIV-status and non-adherence among all APHIV in the study. There was also no association between awareness of status and viral suppression. Most studies have been unable to assess the association between HIV-status awareness and virologic outcomes, instead relying on self-reported ART adherence which may be prone to recall or social desirability bias [9,56]. Our results suggest disclosure may not be associated with long-term non-adherence [27,57], and age-appropriate disclosure should be encouraged to support long-term adherence and positive health trajectories.

The strengths of this study include its large community traced sample which is likely to be representative of APHIV in the study area. The ability to follow participants over three rounds and the high rates of retention enabled longitudinal analysis of changes in HIV-treatment and mental health outcomes. However, our findings should also be interpreted considering the following limitations. First, awareness of status was taken as a dichotomous variable, failing to account for the evolving nature of disclosure. Some adolescents may have had partial knowledge of their status. Second, most of the study variables were assessed by self-report. Even though standardised tools were utilised, there remains potential for social desirability or misclassification bias. Third, the incomplete availability of viral load results and the wide interval between interview dates and results may likely have influenced our results. Lastly, the low prevalence of mental health symptoms meant we were unable to examine clinical levels of mental health problems.

## Conclusions

Older age and residing in an urban location were factors associated with disclosure among adolescents with perinatally-acquired HIV in Eastern Cape, South Africa. There was no increase in mental health symptoms associated with adolescents learning their HIV-positive status. Adolescents who were aware of their HIV-positive status were also not more likely to have higher rates of mental health symptoms, or lower rates of ART adherence or viral suppression. These findings suggest that disclosure is unlikely to have negative HIV-treatment and mental health outcomes among APHIV and should be encouraged alongside post-disclosure adherence support to enable APHIV to take charge of their treatment.

## Data Availability

All data produced in the present study are available upon reasonable request to the authors

## Author’s contributions

OE performed the analysis and wrote the first draft of the paper. YS, JWE and LO provided overall supervision and guidance of statistical analysis and contributed to revisions of the manuscript. LC and ET conceptualized the study on which this manuscript is based, were the lead investigators, and contributed to revisions of the manuscript. SZ contributed to creating the final study dataset, reconstruction of study variables and revisions of the manuscript. All authors have read and approved the final manuscript.

## Acknowledgments

We thank all the young people and their caregiving families who agreed to participate in this study, and the fieldwork team who have shown great dedication and resilience over the years.

## Additional files

Figure S1: Conceptual framework

Table S1. Characteristics of study participants stratified by awareness of their HIV-positive status at the three study rounds

Table S2. Random-intercepts logistic regression results showing odds of learning HIV-positive status (disclosure) between study rounds 1-2 and 2-3 by demographic and psychosocial factors at the survey prior to disclosure (baseline (T1) and second round (T2))

Table S3: Differences in demographic variables between individuals with and without viral load results at all study timepoints.

Table S4. Random-intercepts logistic regression results showing crude and adjusted odds ratio and 95% confidence intervals for factors associated with self-reporting past week ART adherence and having a suppressed viral load within a year before or after interview

Table S5. Random-intercepts logistic regression results showing crude and adjusted odds ratio and 95% confidence intervals for factors associated with self-reporting a symptom of anxiety within the past month, depression within the past two weeks, behavioural problem within the past six months and suicidal idea within the past month of study interview

Table S6. Differential change in the odds of reporting any symptom of anxiety, depression and suicidality or ART adherence between rounds 1 (T1) and 2 (T2), between those who became aware of their HIV-status between surveys, versus those already aware of their status at baseline.

