## Supplementary material for "Predictors and consequences of HIV status disclosure to adolescents living with HIV in Eastern Cape, South Africa"

**Content**

**Figure S1:** Conceptual framework

**Table S1.** Characteristics of study participants stratified by awareness of their HIV-positive status at the three study rounds

**Table S2.** Random-intercepts logistic regression results showing odds of learning HIV-positive status (disclosure) between study rounds 1-2 and 2-3 by demographic and psychosocial factors at the survey prior to disclosure (baseline (T1) and second round (T2))

**Table S3:** Differences in demographic variables between individuals with and without viral load results at all study timepoints.

**Table S4.** Random-intercepts logistic regression results showing crude and adjusted odds ratio and 95% confidence intervals for factors associated with self-reporting past week ART adherence and having a suppressed viral load within a year before or after interview

**Table S5.** Random-intercepts logistic regression results showing crude and adjusted odds ratio and 95% confidence intervals for factors associated with self-reporting a symptom of anxiety within the past month, depression within the past two weeks, behavioural problem within the past six months and suicidal idea within the past month of study interview

**Table S6.** Differential change in the odds of reporting any symptom of anxiety, depression and suicidality or ART adherence between rounds one and two, between those who became aware of their HIV status between surveys, versus those already aware of their status at baseline.

### Figure S1: Conceptual framework

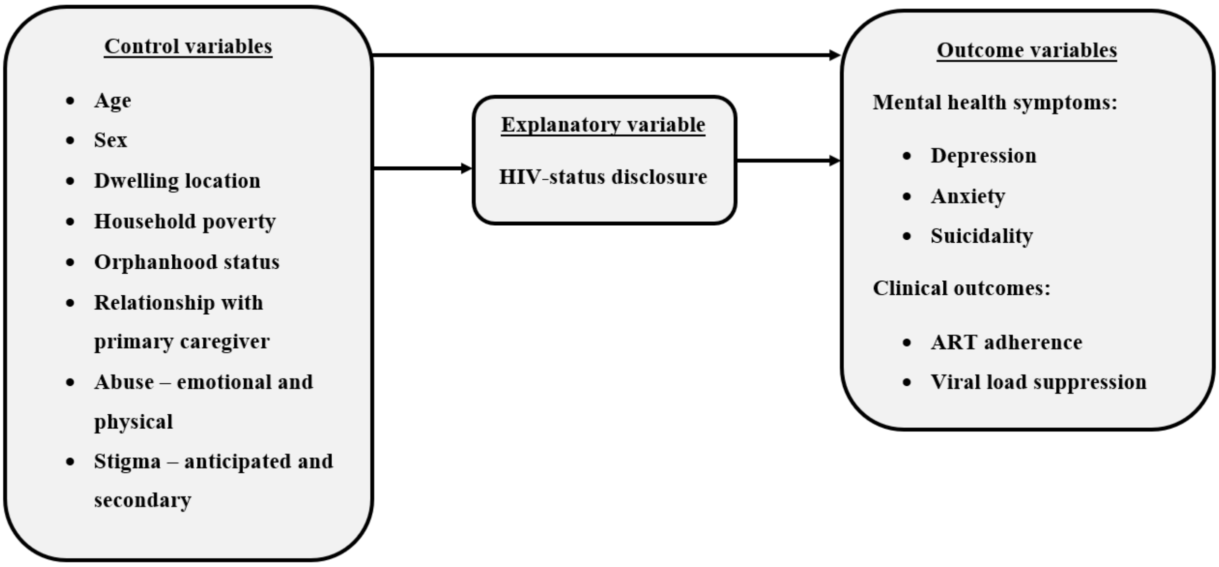

**Table S1. Characteristics of study participants stratified by awareness of their HIV-positive status at the three study rounds**

| | Study round | | | | | | | | |
|---|---|---|---|---|---|---|---|---|---|
| | **Round 1 (N = 813)** | | | **Round 2 (N = 769)** | | | **Round 3 (N = 729)** | | |
| | **Unaware n = 300** | **Aware n = 513** | **p-value** | **Unaware n = 151** | **Aware n = 618** | **p-value** | **Unaware n = 106** | **Aware n = 623** | **p-value** |
| Age in years, mean (SD) | 11.5 (1.76) | 13.6 (2.40) | <0.0001 | 12.5 (1.48) | 14.8 (2.41) | <0.0001 | 13.6 (1.56) | 15.8 (2.45) | <0.0001 |
| Female, n (%) | 149 (49.7) | 261 (50.9) | 0.8 | 71 (47.0) | 319 (51.6) | 0.4 | 51 (48.1) | 318 (51.0) | 0.7 |
| Urban dwelling, n (%) | 203 (67.7) | 408 (79.5) | 0.0002 | 90 (59.6) | 491 (79.6) | <0.0001 | 59 (55.7) | 497 (79.8) | <0.0001 |
| Caregiver is biological parent, n (%) | 149 (49.7) | 199 (38.8) | 0.003 | 61 (40.4) | 235 (38.0) | 0.7 | 44 (41.5) | 233 (37.4) | 0.5 |
| Household poverty, n (%) | 195 (65.0) | 336 (65.5) | 0.9 | 128 (84.8) | 461 (74.6) | 0.01 | 68 (64.2) | 416 (66.8) | 0.7 |
| Any parental loss, n (%) | 146 (48.7) | 356 (69.4) | <0.0001 | 63 (41.7) | 422 (68.3) | <0.0001 | 61 (57.5) | 456 (73.2) | 0.0002 |
| Any emotional abuse in last year, n (%) | 65 (21.7) | 137 (26.7) | 0.1 | 25 (16.6) | 176 (28.5) | 0.004 | 16 (15.1) | 122 (19.6) | 0.3 |
| Any physical abuse in last year, n (%) | 99 (33.0) | 162 (31.6) | 0.7 | 34 (22.5) | 173 (28.0) | 0.2 | 19 (17.9) | 91 (14.6) | 0.5 |
| Any anticipated stigma reported, n (%) | 64 (21.3) | 134 (26.1) | 0.1 | 24 (15.9) | 131 (21.2) | 0.2 | 12 (11.3) | 103 (16.5) | 0.2 |
| Any secondary stigma in last year, n (%) | 45 (15.0) | 105 (20.5) | 0.06 | 9 (6.0) | 54 (8.7) | 0.3 | 1 (0.9) | 26 (4.2) | 0.2 |

| | | | | | | | | | |
|---|---|---|---|---|---|---|---|---|---|
| Self-reported past week ART adherence, n (%) | 197 (65.7) | 362 (70.6) | 0.2 | 107 (70.9) | 410 (66.3) | 0.3 | 88 (83.0) | 480 (77.0) | 0.2 |
| Any depression symptom in past two weeks, n (%) | 126 (42.0) | 226 (44.1) | 0.6 | 43 (28.5) | 217 (35.1) | 0.2 | 33 (31.3) | 172 (27.6) | 0.5 |
| Any anxiety symptom in past month, n (%) | 185 (61.7) | 320 (62.4) | 0.9 | 30 (19.9) | 188 (30.4) | 0.01 | 25 (23.6) | 158 (25.4) | 0.8 |
| Any suicidality symptom in past month, n (%) | 11 (3.7) | 35 (6.8) | 0.09 | 0 (0.0) | 24 (3.9) | 0.03 | 0 (0.0) | 24 (3.9) | 0.08 |

[†] Type of dwelling data missing for 1 (0.1%) adolescent at round 2.

**Table S2. Random-intercepts logistic regression results showing odds of learning HIV-positive status (disclosure) between study rounds 1-2 and 2-3 by demographic and psychosocial factors at the survey prior to disclosure (baseline and second round)**

| | Disclosure status mean (SD) or n (%) | | | | | |
|---|---|---|---|---|---|---|
| | Round 1-2 N = 282 | | Round 2-3 N = 146 | | | |
| **Participant characteristics** | **No disclosure n = 151** | **Disclosure n = 131** | **No disclosure n = 106** | **Disclosure n = 40** | **Odds ratio (95% CI)** | **Adjusted odds ratio (95% CI)** |
| Age | 11.2 (1.36) | 11.9 (1.97) | 12.5 (1.55) | 12.4 (1.21) | 1.19 (1.00 – 1.42)* | 1.24 (1.07 – 1.44)** |
| Female | 71 (47.0) | 71 (54.2) | 51 (48.1) | 17 (42.5) | 1.23 (0.76 – 2.01) | 1.19 (0.78 – 1.82) |
| Urban dwelling | 84 (55.6) | 103 (78.6) | 59 (55.7) | 27 (67.5) | 3.04 (1.72 – 5.36)*** | 2.83 (1.71– 4.67)*** |
| Caregiver is biological parent | 76 (50.3) | 62 (47.3) | 41 (38.7) | 15 (37.5) | 0.96 (0.60 – 1.54) | 0.85 (0.54 – 1.37) |
| Household poverty | 105 (69.5) | 82 (62.6) | 88 (83.0) | 35 (87.5) | 0.72 (0.43 – 1.18) | 0.81 (0.51 – 1.30) |
| Any parental loss | 63 (41.7) | 75 (57.3) | 49 (46.2) | 12 (30.0) | 1.50 (0.91 – 2.47) | 1.11 (0.69 – 1.81) |
| Any emotional abuse in last year | 25 (16.6) | 34 (26.0) | 17 (16.0) | 7 (17.5) | 1.66 (0.94 – 2.92) | 1.21 (0.71 – 2.06) |
| Any physical abuse in last year | 49 (32.5) | 44 (33.6) | 25 (23.6) | 9 (22.5) | 1.10 (0.66 – 1.81) | 0.89 (0.56 – 1.42) |
| Any anticipated stigma reported | 27 (17.9) | 31 (23.7) | 18 (17.0) | 6 (15.0) | 1.38 (0.77 – 2.48) | 1.15 (0.68 – 1.94) |

| | | | | | | |
|---|---|---|---|---|---|---|
| Any secondary stigma in last year | 22 (14.6) | 20 (15.3) | 6 (5.7) | 3 (7.5) | 1.20 (0.60 – 2.43) | 1.00 (0.53 – 1.89) |
| Any depression symptom | 58 (38.4) | 58 (44.3) | 31 (29.2) | 7 (17.5) | 1.11 (0.69 – 1.79) | 0.93 (0.59 – 1.45) |
| Any anxiety symptom | 95 (62.9) | 80 (61.1) | 22 (20.8) | 7 (17.5) | 1.15 (0.72 – 1.82) | 0.80 (0.50 – 1.30) |
| Any suicidality risk | 4 (2.6) | 7 (5.3) | 0 (0.0) | 0 (0.0) | 2.60 (0.62 – 10.95) | 1.80 (0.46 – 7.10) |
| Round 2 | 151 (53.6) | 131 (46.4) | - | - | Ref | Ref |
| Round 3 | - | - | 106 (72.6) | 40 (27.4) | | 0.37 (0.22 – 0.62)*** |

***p ≤ 0.001, **p ≤ 0.01, *p ≤ 0.05; All models were adjusted for age, sex, dwelling type, caregiver relationship, orphanhood status, anticipated and secondary stigma, and study round.

**Table S3: Differences in demographic variables between individuals with and without viral load results at all study timepoints.**

| | Round 1 (N = 813) Mean(SD) or n (%) | | | Round 2 (N = 769) Mean(SD) or n (%) | | | Round 3 (N = 729) Mean(SD) or n (%) | | |
|---|---|---|---|---|---|---|---|---|---|
| | **With result (n = 508)** | **Without result (n = 305)** | **p-value** | **With result (n = 466)** | **Without result (n = 303)** | **p-value** | **With result (n = 287)** | **Without result (n = 442)** | **p-value** |
| Age in years | 12.7 (2.37) | 13.1 (2.43) | 0.01 | 14.1 (2.31) | 14.8 (2.57) | <0.0001 | 15.1 (2.22) | 15.7 (2.58) | 0.004 |
| Female | 256 (50.4) | 154 (50.5) | 1 | 229 (49.1) | 161 (53.1) | 0.3 | 133 (46.3) | 236 (53.4) | 0.07 |
| Aware of HIV status | 318 (62.6) | 195 (63.9) | 0.8 | 364 (78.1) | 254 (83.8) | 0.06 | 242 (84.3) | 381 (86.2) | 0.6 |
| Caregiver is biological parent | 225 (44.3) | 123 (40.3) | 0.3 | 184 (39.5) | 112 (37.0) | 0.5 | 112 (39.0) | 165 (37.3) | 0.7 |

**Table S4. Random-intercepts logistic regression results showing crude and adjusted odds ratio and 95% confidence intervals (CI) for factors associated with self-reporting past week ART adherence and having a suppressed viral load within a year before or after interview**

| | Self-reported past week ART adherence (N = 729) | | Viral suppression (N = 453, 444 and 287 at rounds 1, 2 and 3) | |
|---|---|---|---|---|
| | Odds ratio (95% CI) | Adjusted odds ratio (95% CI) | Odds ratio (95% CI) | Adjusted odds ratio (95% CI)) |
| **Explanatory variable** | | | | |
| Aware of HIV status | 1.04 (0.82 – 1.32) | 1.03 (0.78 – 1.36) | 0.30 (0.10 – 0.90)* | 0.81 (0.22 – 2.98) |
| **Control variables** | | | | |
| Age | 1.00 (0.96 – 1.04) | 0.96 (0.92 – 1.01) | 0.65 (0.52 – 0.81)*** | 0.92 (0.69 – 1.24) |
| Female | 1.03 (0.83 – 1.27) | 1.04 (0.83 – 1.29) | 0.93 (0.30 – 2.93) | 0.84 (0.22 – 3.24) |
| Urban dwelling | 1.03 (0.81 – 1.32) | 1.13 (0.87 – 1.45) | 0.52 (0.14 – 1.92) | 0.47 (0.10 – 2.19) |
| Caregiver is biological parent | 0.83 (0.67 – 1.02) | 0.82 (0.64 – 1.04) | 1.28 (0.54 – 3.00) | 1.40 (0.46 – 4.19) |
| Household poverty | 0.96 (0.77 – 1.19) | 0.92 (0.74 – 1.15) | 0.50 (0.23 – 1.11) | 0.43 (0.17 – 1.11) |
| Any parental loss | 1.13 (0.91 – 1.40) | 1.04 (0.81 – 1.35) | 0.69 (0.26 – 1.87) | 1.63 (0.46 – 5.81) |
| Any emotional abuse in last year | 0.53 (0.42 – 0.66)*** | 0.65 (0.51 – 0.84)*** | 2.85 (1.07 – 7.59)* | 3.46 (1.01 – 11.89)* |
| Any physical abuse in last year | 0.54 (0.43 – 0.68)*** | 0.68 (0.53 – 0.87)** | 2.54 (1.05 – 6.19)* | 1.28 (0.43 – 3.83) |
| Any anticipated stigma reported | 1.82 (1.53 – 2.34)** | 0.85 (0.66 – 1.10) | 2.67 (0.98 – 7.26) | 1.80 (0.58 – 5.57) |
| Any secondary stigma in last year | 0.58 (0.42 – 0.79)*** | 0.77 (0.55 – 1.08) | 0.85 (0.29 – 2.54) | 0.28 (0.07 – 1.08) |
| Round 2 | 0.93 (0.73 – 1.16) | 0.97 (0.76 – 1.24) | 0.61 (0.28 – 1.39) | 0.75 (0.28 – 2.02) |
| Round 3 | 1.63 (1.28 – 2.08)*** | 1.63 (1.24 – 2.16)*** | 0.08 (0.03 – 0.20)*** | 0.09 (0.02 – 0.31)*** |

***p ≤ 0.001, **p ≤ 0.01, *p ≤ 0.05

**Table S5. Random-intercepts logistic regression results showing crude and adjusted odds ratios (OR) and 95% confidence intervals for factors associated with self-reporting a symptom of anxiety within the past month, depression within the past two weeks and suicidal idea within the past month of study interview**

| | Anxiety | | Depression | | Suicidality | |
|---|---|---|---|---|---|---|
| **(N = 729)** | **Odds ratio (95% CI)** | **Adjusted odds ratio (95% CI)** | **Odds ratio (95% CI)** | **Adjusted odds ratio (95% CI)** | **Odds ratio (95% CI)** | **Adjusted odds ratio (95% CI)** |
| **Explanatory variable** | | | | | | |
| Aware of HIV status | 0.79 (0.65 – 0.97)* | 1.03 (0.79 – 1.35) | 0.97 (0.78 – 1.20) | 0.89 (0.69 – 1.13) | 1.07 (0.40 – 2.84) | 1.42 (0.42 – 4.75) |
| **Control variables** | | | | | | |
| Age | 0.92 (0.89 – 0.96)*** | 1.02 (0.97 – 1.07) | 1.02 (0.99 – 1.06) | 1.07 (1.03 – 1.12)** | 0.98 (0.82 – 1.17) | 1.21 (0.96 – 1.52) |
| Female | 1.15 (0.97 – 1.36) | 1.19 (0.97 – 1.46) | 0.99 (0.83 – 1.20) | 0.98 (0.81 – 1.18) | 1.97 (0.68 – 5.65) | 1.88 (0.62 – 5.69) |
| Urban dwelling | 1.00 (0.82 – 1.22) | 0.89 (0.70 – 1.13) | 1.08 (0.87 – 1.34) | 1.03 (0.82 – 1.28) | 1.09 (0.36 – 3.30) | 0.73 (0.23 – 2.32) |
| Caregiver is biological parent | 1.05 (0.88 – 1.25) | 0.97 (0.77 – 1.23) | 0.98 (0.81 – 1.18) | 0.97 (0.78 – 1.21) | 0.86 (0.36 – 2.12) | 0.79 (0.27 – 2.26) |
| Household poverty | 0.82 (0.68 – 0.99)* | 0.92 (0.74 – 1.14) | 0.96 (0.78 – 1.17) | 0.97 (0.79 – 1.19) | 1.38 (0.64 – 2.98) | 1.21 (0.53 – 2.73) |

| | | | | | | |
|---|---|---|---|---|---|---|
| Any parental loss | 0.95 (0.79 – 1.14) | 0.98 (0.77 – 1.26) | 1.03 (0.85 – 1.25) | 0.97 (0.77 – 1.22) | 0.83 (0.32 – 2.17) | 0.76 (0.24 – 2.40) |
| Any emotional abuse in last year | 2.95 (2.40 – 3.62)*** | 2.34 (1.82 – 3.02)*** | 2.25 (1.82 – 2.78)*** | 1.80 (1.43 – 2.27)*** | 4.95 (2.43 – 10.07)*** | 2.57 (1.09 – 6.04)** |
| Any physical abuse in last year | 2.71 (2.21 – 3.33)*** | 1.86 (1.46 – 2.38)*** | 1.59 (1.29 – 1.96)*** | 1.12 (0.89 – 1.41) | 3.84 (1.86 – 7.95)*** | 2.10 (0.88 – 5.04) |
| Any anticipated stigma reported | 1.86 (1.50 – 2.29)*** | 1.38 (1.08 – 1.78)* | 1.78 (1.43 – 2.22)*** | 1.37 (1.09 – 1.73)** | 4.96 (2.33 – 10.59)*** | 2.87 (1.28 – 6.45)* |
| Any secondary stigma in last year | 5.61 (4.08 – 7.72)*** | 3.24 (2.26 – 4.64)*** | 3.44 (2.57 – 4.59)*** | 2.42 (1.78 – 3.29)*** | 12.16 (5.15 – 28.68)*** | 6.46 (2.59 – 16.16)*** |
| Round 2 | 0.22 (0.17 – 0.28)*** | 0.21 (0.16 – 0.28)*** | 0.66 (0.53 – 0.82)*** | 0.66 (0.52 – 0.84)*** | 0.31 (0.15 – 0.67)** | 0.29 (0.12 – 0.70)** |
| Round 3 | 0.18 (0.14 – 0.23)*** | 0.21 (0.16 – 0.28)*** | 0.51 (0.41 – 0.64)*** | 0.54 (0.41 – 0.69)*** | 0.36 (0.17 – 0.77)** | 0.44 (0.16 – 1.20) |

***$p \leq 0.001$, **$p \leq 0.01$, *$p \leq 0.05$

**Table S6. Differential change in the odds of reporting any symptom of anxiety, depression and suicidality or ART adherence between rounds 1 (R1) and 2 (R2), between those who became aware of their HIV status between surveys, versus those already aware of their status at baseline.**

| | n (%) | | | | †Differential change estimate (95%CI) | |
|---|---|---|---|---|---|---|
| | Aware at baseline (n = 487) | | Disclosure R1-R2 (n = 131) | | | |
| | **Round 1** | **Round 2** | **Round 1** | **Round 2** | **Crude** | **‡Adjusted** |
| **Anxiety** | 304 (62.4) | 147 (30.2) | 80 (61.1) | 41 (31.3) | 1.12 (0.62 – 1.97) | 1.14 (0.63 – 2.04) |
| **Depression** | 211 (43.3) | 173 (35.5) | 58 (44.3) | 44 (33.6) | 0.88 (0.50 – 1.55) | 0.88 (0.49 – 1.56) |
| **Suicidality** | 31 (6.4) | 19 (3.9) | 7 (5.3) | 5 (3.8) | 1.18 (0.30 – 4.35) | 1.20 (0.30 – 4.64) |
| **ART adherence** | 346 (71.0) | 333 (68.3) | 92 (70.2) | 77 (58.7) | 0.69 (0.38 – 1.22) | 0.68 (0.38 – 1.22) |

***p ≤ 0.001, **p ≤ 0.01, *p ≤ 0.05. †The differential change estimate represents the ratio of the odds ratios of reporting any symptom of anxiety, depression and suicidality or ART adherence between R1 and R2 between those who became aware vs. those remaining unaware.

‡Adjusted for age, sex, dwelling location, household poverty, caregiver relationship, orphanhood status, physical and emotional abuse, and anticipated and secondary stigma.
